# Inhibitory KIR ligands are associated with higher *P. falciparum* parasite prevalence

**DOI:** 10.1101/2020.04.04.20053504

**Authors:** Jean C. Digitale, Perri C. Callaway, Maureen Martin, George Nelson, Mathias Viard, John Rek, Emmanuel Arinaitwe, Grant Dorsey, Moses Kamya, Mary Carrington, Isabel Rodriguez-Barraquer, Margaret E. Feeney

**Author notes:** Correspondence: Margaret Feeney, 1001 Potrero Avenue, Building 3, Room 545a, San Francisco CA 94110. These authors contributed equally (Jean C. Digitale, Perri C. Callaway, Isabel Rodriguez-Barraquer, Margaret E. Feeney). The authors have declared that no conflict of interest exists.

## Abstract

Killer cell immunoglobulin-like receptors (KIR) and their HLA ligands influence the outcome of many infectious diseases. We analyzed the relationship of compound KIR-HLA genotypes with risk of *Plasmodium falciparum* infection in a longitudinal cohort of 890 Ugandan individuals. We found that presence of HLA-C2 and HLA-Bw4, ligands for inhibitory KIR2DL1 and KIR3DL1, respectively, increased the likelihood of *P. falciparum* parasitemia in an additive manner. Individuals homozygous for HLA-C2, which mediates strong inhibition via KIR2DL1, had the highest odds of parasitemia, HLA-C1/C2 heterozygotes had intermediate odds, and individuals homozygous for HLA-C1, which mediates weaker inhibition through KIR2DL2/3, had the lowest odds of parasitemia. Additionally, higher surface expression of HLA-C, the ligand for inhibitory KIR2DL1/2/3, was associated with a higher likelihood of parasitemia. Together these data indicate that stronger KIR-mediated inhibition confers a higher risk of *P. falciparum* parasitemia and suggest that KIR-expressing effector cells play a role in mediating anti-parasite immunity.

## Introduction

*Plasmodium falciparum* malaria has caused hundreds of millions of deaths throughout history and continues to kill nearly half of a million people each year (World Health Organization, 2018). As a result, *P. falciparum* has exerted strong selective pressure on human evolution (Kariuki and Williams, 2020; Kwiatkowski, 2005). Most of the human genetic mutations that are known to alter malaria susceptibility lead to erythrocyte defects or cytoadhesion variants (Kariuki and Williams, 2020; Kwiatkowski, 2005). However, a recent GWAS analysis of 17,000 individuals estimated that known variants account for only 11% of the total host genetic influence on malaria susceptibility (Band et al., 2019). Surprisingly, only a few gene variants implicated in the host immune response have been robustly associated with malaria susceptibility. Such associations could be particularly valuable in illuminating causal mechanisms of immune protection. Notably, most studies of host genetic susceptibility to malaria, including the GWAS study noted above, have compared severe malaria cases to controls with non-severe (but symptomatic) malaria (Hirayasu et al., 2012; Lourembam et al., 2011). This analytic framework could overlook genes that influence protection at the earlier pre-erythrocytic stages of the *P. falciparum* life cycle, before blood stage infection is established.

Human killer cell immunoglobulin-like receptors (KIR) are a family of diverse inhibitory and activating transmembrane receptors whose known ligands are primarily HLA class I molecules. The KIR locus is the second most polymorphic region in the human genome after HLA, and is evolving even more rapidly, likely in part due to selective pressure from pathogens (Kulkarni et al., 2008). Human haplotypes vary both in the number of KIR genes and in the proportion that are inhibitory or activating. While some KIR genes are universal, others are expressed by only a subset of individuals (Middleton and Gonzelez, 2010). Moreover, within an individual, only a fraction of cells express a given KIR molecule (Andersson et al., 2009). KIR are expressed on natural killer (NK) cells, some gamma delta T cells, and some CD8 T cells, all of which play a role in the antimalarial immune response (Burrack et al., 2019; Dantzler and Jagannathan, 2018; Lefebvre and Harty, 2019). Interactions of KIR with their HLA ligands modulate the activation threshold of the cell and have been shown to influence the outcome of several, predominantly viral, infections (Bona et al., 2017; Khakoo et al., 2004; Mori et al., 2019). Because the KIR and HLA loci are on different chromosomes and segregate independently, associations of the KIR-HLA compound genotype with susceptibility to a particular pathogen can shed light on mechanisms critical for immunity and immunopathogenesis.

To date, the association of KIR and their HLA ligands with susceptibility to malaria has been examined in very few studies and almost entirely in the context of case-control comparisons of severe vs. non-severe malaria (Hirayasu et al., 2012; Lourembam et al., 2011). Here, we examined the influence of well-defined KIR-HLA ligand pairs on the prevalence of *P. falciparum* parasitemia and malaria in a longitudinal cohort of 890 individuals from three sites with varying malaria transmission in Uganda to determine the influence of KIR on susceptibility to *P. falciparum* infection. This analysis revealed that individuals with more inhibitory KIR-HLA pairs have higher odds of parasitemia, strongly supporting a role for cellular immunity in host restriction of the parasite.

## Results

### Clinical cohort and KIR-HLA genotyping

We analyzed data from 657 children and 233 adults recruited from 292 households at three study sites in Uganda (Table 1). Genotyping of KIR and HLA class I loci was performed (Table 2), and individuals were categorized based on the presence of well-characterized KIR ligand groups (i.e. HLA-C2 for KIR2DL1, HLA-C1 for KIR2DL2/3, and HLA-Bw4 for KIR3DL1, Supplemental Table 1). The prevalence of the inhibitory KIR genes KIR2DL1 and KIR3DL1 was very high (both >98.5%), consistent with previous reports in East African populations (Nakimuli et al., 2013). In order to control for KIR-HLA differences in ethnicity, participants’ ethnicity was categorized as Bantu or non-Bantu based on the language of their consent form (Table 1). However, as ethnicity was largely collinear with site, including it in models did not improve model fit beyond site fixed effects; it was therefore omitted from final analyses.

**Table 1.** Characteristics of study population.

|  | **Jinja** | **Kanungu** | **Tororo** |
| --- | --- | --- | --- |
| **Households (N)** | **97** | **100** | **95** |
| Median household EIR (range) | 2.1 (1.5-22.0) | 6.3 (3.0-44.5) | 67.6 (11.5-588.4) |
| **Adults (N)** | **89** | **78** | **66** |
| Median incidence of symptomatic malaria episodes per person year (range) | 0.0 (0.0-2.0) | 0.2 (0.0-3.7) | 0.0 (0.0-3.0) |
| Parasite prevalence (% quarters parasite+) | 4.9% | 12.8% | 12.0% |
| **Children (N)** | **187** | **267** | **203** |
| Median age at enrollment, years (range) | 4.4 (0.5-10.0) | 4.7 (0.5-9.9) | 5.1 (0.6-10.0) |
| Median incidence of symptomatic malaria episodes per person year (range) | 0.2 (0.0-5.1) | 1.2 (0.0-8.1) | 2.1 (0.0-8.8) |
| Parasite prevalence (% quarters parasite +, range) | 12.8% | 36.4% | 64.2% |
| **Median follow up time, months (range)\*** | **55.1 (2.3-58.6)** | **56.7 (0.0-58.3)** | **39.0 (1.8-40.3)** |
| **Sickle cell, n(%)** |  |  |  |
| AA | 201 (72.8%) | 323 (93.6%) | 195 (72.5%) |
| AS | 72 (26.1%) | 22 (6.4%) | 70 (26.0%) |
| SS | 0 (0.0%) | 0 (0.0%) | 2 (0.7%) |
| No result | 3 (1.1%) | 0 (0.0%) | 2 (0.7%) |
| **Bantu ethnicity, N (%)** | **180 (65.5%)** | **341 (98.8%)** | **2 (0.7%)** |

\*Note: 3 people in the sample had only one visit; Tororo visits after December 31, 2014 were excluded because transmission changed dramatically at this site after intensive indoor residual spraying campaigns were started.

**Table 2.** KIR and Ligand Prevalence.

| KIR and HLA prevalence | N=890 |
| --- | --- |
| KIR | % |
| KIR2DL1† | 98.6 |
| KIR2DL2 | 54.7 |
| KIR2DL3‡ | 84.8 |
| KIR2DL4 | 99.7 |
| KIR2DL5 | 57.9 |
| KIR2DP1 | 98.1 |
| KIR2DS1 | 23.5 |
| KIR2DS2 | 49.0 |
| KIR2DS3 | 23.0 |
| KIR2DS4 | 68.3 |
| KIR2DS5 | 42.6 |
| KIR3DL1 | 98.7 |
| KIR3DL2 | 99.6 |
| KIR3DS1 | 13.0 |
| HLA | % |
| C1/C2 ligand group zygosity* |  |
| C1/C1 | 19.5 |
| C1/C2 | 53.1 |
| C2/C2 | 27.4 |
| Bw4 ligand group (at least one copy) | 74.8 |

Note: Reduced Ns due to failed amplification or lack of DNA.
†N=888
‡N=883
\*N=889

Subjects were followed longitudinally from August 2011-June 2016 with blood smears performed every 3 months to assess *P. falciparum* parasitemia. Reflecting the varying transmission intensity among the sites, the prevalence of parasitemia was highest among children in Tororo (64%), followed by Kanungu (36%) and Jinja (13%). The incidence of malaria, defined as parasitemia accompanied by fever (presented at clinic with temperature >38.0 or self-reported in last 24 hours), also differed among the three sites and was inversely associated with age (Rodriguez-Barraquer et al., 2018). Both malaria incidence and parasite prevalence by microscopy were substantially lower among adults than children (Table 1).

To assess whether KIR and their HLA class I ligands influence the risk of *P. falciparum* infection or its clinical manifestations, we tested associations of individual KIR genes, HLA ligand groups and KIR-HLA compound genotypes with 5 malaria outcome measures: prevalence of parasitemia, clinical malaria incidence, parasite density (stratified by routine quarterly visits and malaria visits), conditional probability of clinical malaria given the presence of parasitemia, and temperature at parasitemic visits, conditional on parasite density. For each association, we report the p-value from a multi-level model (p), an empirical p-value after permutation tests (p*), and a q-value (q) calculated by applying the false discovery rate correction for multiple comparisons to the empirical p-values as described in Methods. There was no association between the presence of any individual KIR gene with any of these outcomes (Supplemental Tables 2a-f), so we next assessed the presence of KIRs in conjunction with their HLA ligands.

### *Inhibitory KIR2DL1 ligand HLA-C2 increases risk of* P. falciparum *parasitemia*

Among the best characterized KIR-HLA ligand interactions are the inhibitory KIR2DL1 with the HLA-C2 subgroup of HLA-C alleles and KIR2DL2/L3 with the HLA-C1 subgroup. HLA-C2 alleles possess a lysine at amino acid position 80 whereas HLA-C1 alleles have an asparagine (Biassoni et al., 1995). Since KIR2DL1 (98.6%) and KIR2DL2/3 (99.3%) were both present in virtually all subjects, the presence of their HLA ligands determines which inhibitory KIR/HLA-C pair(s) an individual has. Further, within these inhibitory KIR/HLA-C ligand interactions, there is a hierarchy of inhibition. KIR2DL1/HLA-C2 provides the strongest inhibition, followed by KIR2DL2/HLA-C1, while KIR2DL3/HLA-C1 provides the weakest inhibition (Carrington et al., 2005; Moesta et al., 2008; Winter et al., 1998). We found that individuals with at least one copy of the KIR2DL1 ligand HLA-C2 had 1.31-fold higher odds of being parasitemic (p=0.017, p*=0.010, q=0.047). Conversely, having at least one HLA-C1 allele decreased the odds of being parasitemic (OR=0.79, p=0.016, p*=0.013, q=0.051).

Next, we assessed the zygosity of HLA-C ligand groups to determine whether the number of copies of either HLA-C1 or HLA-C2 influenced parasite prevalence. HLA-C1/C1 individuals are only able to receive inhibitory signals via KIR2DL2/3, whereas HLA-C2/C2 individuals receive inhibitory signals predominantly via KIR2DL1, and heterozygous individuals have the ligand for all three of these KIRs (Biassoni et al., 1995). Comparison of these groups revealed a stepwise increase in parasite prevalence, with HLA-C2/C2 individuals having 1.54-fold higher odds of parasitemia compared to HLA-C1/C1 individuals (Table 3). Heterozygous (HLA-C1/C2) individuals had intermediate odds. These results suggest that engagement of KIR2DL1 with its ligand HLA-C2, the most inhibitory of the KIR/HLA-C ligand pairs, increases parasite prevalence in comparison to engagement of KIR2DL2/3 with their ligand HLA-C1, which in general provide weaker inhibitory signals. KIR2DL1 expressing cells from HLA-C2/C2 homozygous individuals will receive stronger inhibitory signals, potentially dampening the effector cell response to *P. falciparum* infection.

**Table 3.** Effect of HLA-C zygosity on parasite prevalence.

| C1/C2 zygosity | OR | P-value | Empirical p-value | Empirical FDR q-value | N |
| --- | --- | --- | --- | --- | --- |
| C1/C1 | Ref |  |  |  | 173 |
| C1/C2 | 1.26 | 0.045 | 0.036 | 0.111 | 472 |
| C2/C2 | 1.54 | 0.001 | 0.001 | 0.008 | 244 |

KIR2DL2 and KIR2DL3 segregate as alleles of the same locus, but KIR2DL2 provides stronger effector cell inhibition when bound to HLA-C1 (Moesta et al., 2008). Overall, no difference in parasitemia risk was observed between KIR2DL3/C1 and KIR2DL2/C1 individuals (Table 4). However, KIR2DL2 is in linkage disequilibrium with the homologous activating receptor KIR2DS2 (Wn*=0.85, Fisher’s exact test p<0.001), which may be capable of binding HLA-C1 ligands in a peptide dependent manner (Moesta and Parham, 2012; Naiyer et al., 2017). To test whether KIR2DS2 may have an opposing effect that could obscure the stronger inhibitory impact of KIR2DL2, we examined the impact of KIR2DL2 and KIR2DL3 among individuals lacking the KIR2DS2 gene. This revealed that presence of at least one copy of KIR2DL2 increased the odds of being parasitemic 1.6-fold compared to those with only KIR2DL3 (Table 4). While KIR2DL3/HLA-C1, the least inhibitory KIR-HLA combination in the hierarchy, was associated with a lower prevalence of *P. falciparum* parasitemia, previous data indicate that it markedly increases the risk of cerebral malaria, an inflammatory complication of malaria (Hirayasu et al., 2012). Together these data further support a hierarchy of inhibition, in which KIR-HLA combinations with stronger inhibitory potential increase parasite prevalence.

**Table 4.**
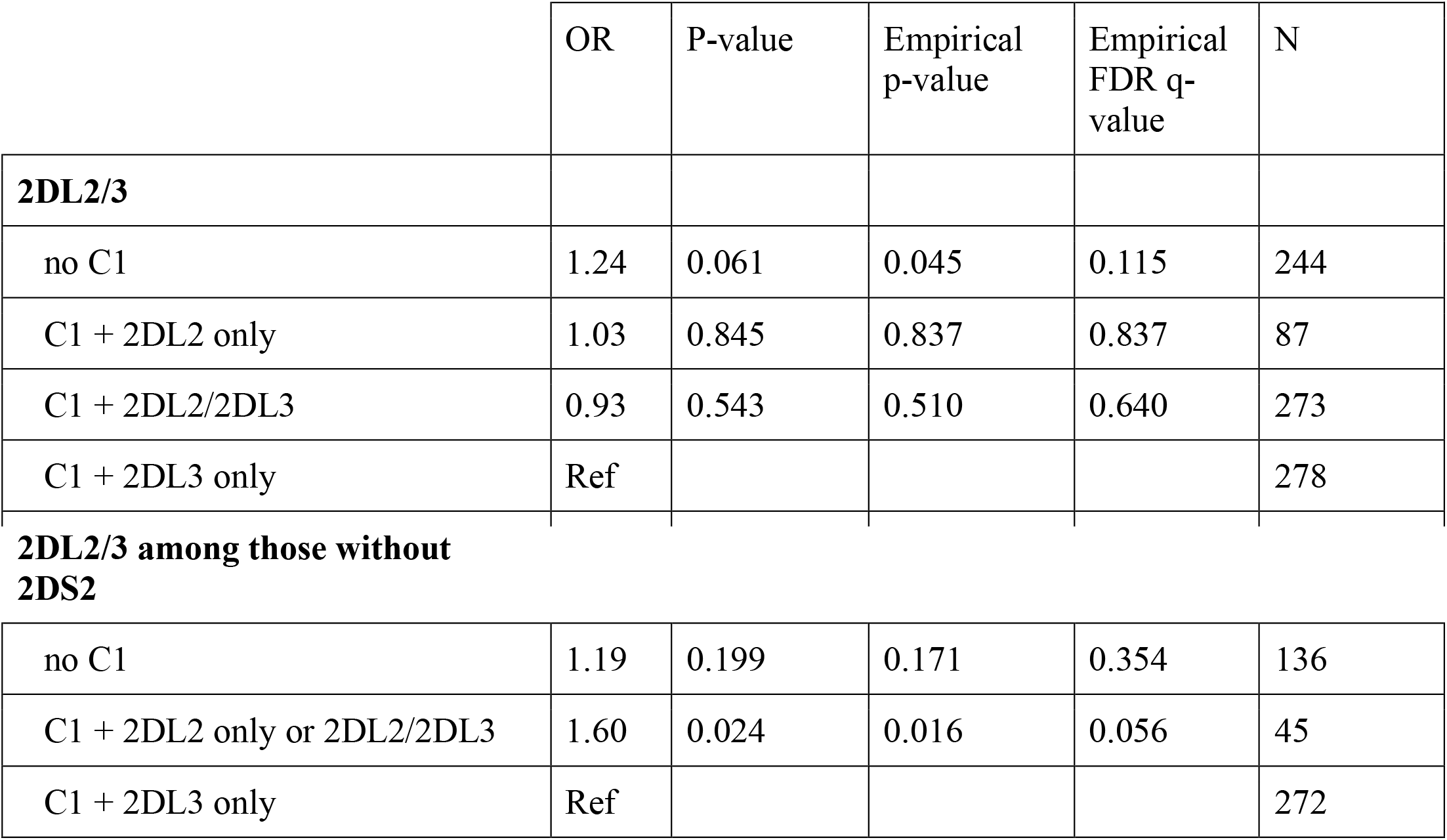
Effect of KIR2DL2/3 on parasite prevalence.

### *Inhibitory KIR3DL1 ligand HLA-Bw4 increases risk of* P. falciparum *parasitemia*

Another inhibitory KIR molecule, KIR3DL1, recognizes a subset of HLA-A and HLA-B alleles that contain the HLA-Bw4 motif, defined by amino acid residues 77-83 on the alpha1-helix (Gumperz et al., 1995). Similar to KIR2DL1, the KIR3DL1 gene was nearly universal in our Ugandan cohort (98.7%), hence its effect on parasite prevalence could not be estimated directly. However, only 74.8% of individuals had an HLA-Bw4 ligand, enabling us to estimate the effect of the KIR-HLA ligand pair. Among individuals with the HLA-Bw4 ligand, the odds of parasitemia were 1.31-fold higher than in individuals lacking HLA-Bw4 (p=0.012, p*=0.006, q=0.040). This further supports the hypothesis that greater inhibition via KIR increases the likelihood of parasitemia, perhaps due to dampened cellular immunity.

Because both HLA-C2 and HLA-Bw4 were found to increase parasite prevalence and they have previously been shown to be in linkage disequilibrium, we looked for an additive effect of the two ligand groups by analyzing the combined genotype. Having both HLA-C2 and HLA-Bw4 increased the odds of being parasitemic by 1.46 times compared to HLA-C2 alone (p=0.001, p*<0.001, q=0.008) and by 1.51 times compared to HLA-Bw4 alone (p=0.002, p*=0.001, q=0.012), suggesting an additive inhibitory effect of the two KIR-HLA ligand pairs. Together, this supports a model wherein the more inhibitory signals that KIR expressing effector cells can receive, the greater the risk of parasitemia.

### Higher surface expression of inhibitory KIR ligand HLA-C is associated with increased odds of parasitemia

To further elucidate the means by which HLA-C influences *P. falciparum* infection, we examined the relationship of HLA-C surface expression levels to *P. falciparum* parasite prevalence. HLA-C molecules have been ascribed two distinct roles in cellular immunity – as ligands for inhibitory KIR, and as antigen presenting molecules for CD8 T cells – both of which can influence infectious disease outcomes (Bona et al., 2017; Khakoo et al., 2004; Marco et al., 2017). In contrast to HLA-A and -B molecules, surface expression of HLA-C is 10-15 fold lower and varies substantially amongst individual allotypes (Apps et al., 2015). During viral infections such as HIV infection, HLA-C alleles that are more highly expressed at the cell surface have been associated with a more potent CD8 T cell response that correlates with enhanced viral control (Corrah et al., 2011; Thomas et al., 2009). We hypothesized that if the protection associated with certain HLA-C ligand groups is attributable to differences in peptide presentation, then higher HLA-C expression would similarly correlate with a reduced likelihood of parasitemia. We assigned each HLA-C allele a value based on its previously determined expression levels (Apps et al., 2013), and calculated a total HLA-C expression level for each individual. We found that higher HLA-C expression was associated with a statistically significant *increase* in the odds of *P. falciparum* parasitemia (1.12-fold increase per one standard deviation of expression; p=0.014, p*=0.006, q=0.040). This contrasts with HIV infection, where highly expressed HLA-C allotypes are protective against HIV. These data suggest that the mechanism by which HLA-C molecules influence restriction of *P. falciparum* parasitemia is not antigen presentation to T cells, but is more consistent with a role in KIR-mediated inhibition of effector cells.

### Relationship of KIR-HLA genotype to other parasitological and clinical malaria outcomes

Finally, because the immune mechanisms responsible for restricting parasitemia may differ from those that protect against symptomatic malaria, we also examined the association of KIR-HLA ligand pairs with the other parasitological and clinical outcomes noted above (Supplemental Table 2b-f). We did not observe an association of KIR-HLA ligand pairings with any of these outcomes. Together these results are most consistent with an influence of KIR-expressing effector cells on restriction of parasite replication, rather than on the downstream inflammatory consequences that eventuate in fever and other clinical symptoms.

In addition, we performed multiple sensitivity analyses to assess the robustness of our results. A more sensitive measure of parasitemia (loop-mediated isothermal amplification—LAMP) was performed on samples that were negative by microscopy at routine visits. However, these data were not included in the primary analyses because LAMP was not conducted consistently on samples from adults at the three sites throughout the study duration. When the definition of parasitemia was expanded to also include the submicroscopic *P. falciparum* infections detectable by LAMP among children for the entire follow-up period (Supplemental Table 3) and among children and adults for the period LAMP data were available for both (Supplemental Table 4), similar associations for all results stated above were observed. In addition, we analyzed models: 1) stratified by site (Supplemental Table 5a-c) to determine whether our results were robust across transmission intensities and ethnicities, 2) restricted to children as they suffer the largest burden of parasitemia (Supplemental Table 6), and 3) excluding individuals with hemoglobin S (sickle cell) mutations to ensure our results were not confounded by sickle cell variants (Supplemental Table 7). These secondary analyses yielded similar results, further strengthening our findings.

## Discussion

Using longitudinal data from Ugandan individuals with varying malaria exposure, we found that the major inhibitory KIR ligands HLA-C2 and HLA-Bw4 were both associated with a higher prevalence of *P. falciparum* parasitemia. Our data indicate that both the number of inhibitory KIR-HLA ligand pairs and the strength of their cellular inhibitory signaling influence the risk of *P. falciparum* parasitemia, but notably, do not impact clinical malaria. Our findings suggest that the cellular immune response plays a critical role either in the prevention or clearance of blood-stage *P. falciparum* infection. These associations represent a novel genetic determinant of *P. falciparum* parasitemia, and support an expanded role for KIR in modulating host immunity to parasitic infections.

This novel association of KIR-HLA compound genotypes with the prevalence of parasitemia is somewhat paradoxical given that *P. falciparum* spends most of its life cycle outside of HLA-expressing cells. Following intradermal inoculation by mosquitoes, *P. falciparum* sporozoites migrate via the dermis and lymphatics to infect hepatocytes, the only stage in the parasite replicative life cycle in which it is within an HLA-expressing host cell (Mota et al., 2001).

Within the hepatocyte, parasites expand 2,000-40,000 fold, then emerge synchronously as merozoites, which establish a self-propagating cycle of erythrocytic infection (Crutcher and Hoffman, 1996; Sturm et al., 2006). Because erythrocyte membranes contain little to no HLA (Moras et al., 2017), we hypothesize that the influence of KIR on cell-mediated anti-parasite immunity may occur primarily during the liver stage when the multiplicity of infection is also lowest. It has previously been shown that increasing KIR mediated NK cell inhibition is associated with poorer clearance of HCV and HBV in the liver (Bona et al., 2017; Khakoo et al., 2004). However, it is also possible that the observed impacts of KIR-HLA pairs on parasite prevalence may be due to enhanced spontaneous clearance of parasitemia rather than cessation of infection at the liver stage. During the blood stage, KIR-expressing effector cells may respond more strongly to an HLA-devoid cell due to the loss of tonic inhibitory signaling via inhibitory KIR (Kim et al., 2008). KIR inhibition may also influence the clearance of parasites through antibody dependent cellular cytotoxicity (ADCC) (Lisovsky et al., 2019; Parsons et al., 2010).

It is notable that we did not observe an influence of KIR or their HLA ligands on symptomatic malaria. In endemic regions, “clinical immunity”—defined as protection from symptomatic malaria–develops gradually with increasing age and exposure to the *P. falciparum* parasite, but sterilizing immunity (i.e. protection against parasitemia) does not (Tran et al., 2013). Hence, it is likely that the immune mechanisms underlying sterile protection and restriction of parasite replication differ from those that mediate clinical immunity, with the latter involving immunoregulatory processes that dampen the inflammation responsible for fever and other symptoms, while fostering successful long-term parasitism. By definition, sterile immunity is directed at the pre-erythrocytic (sporozoite and liver) stages of parasite development. The pre-erythrocytic immune response can prevent or greatly limit the establishment of blood-stage parasitemia and is believed to be mediated by CD8 T cells (Hafalla et al., 2006; Lefebvre and Harty, 2019), NK cells (Arora et al., 2018; Hart et al., 2019, 2017) and the semi-innate V*γ*9Vδ2 subset of *γ*δ T cells (Farrington et al., 2016; Jagannathan et al., 2014; Mamedov et al., 2018; Zaidi et al., 2017), all of which can express KIR. It remains possible that KIR-HLA interactions exert an impact on the symptomatic or inflammatory manifestations of malaria that we were underpowered to detect, given that all members of this study cohort had access to free clinical care which afforded prompt diagnosis and treatment of malaria. Indeed, Hirayasu et. al. reported that the risk of cerebral malaria, an inflammatory complication, is markedly elevated among individuals with KIR2DL3/HLA-C1, which is the least inhibitory pairing amongst KIR2DL1/2/3 ligands (Hirayasu et al., 2012). Immunopathology contributes substantially to the neurological damage caused by cerebral malaria (Grau et al., 1989) and NK cells have been found to accumulate in the brain microvasculature (Hansen et al., 2007). Thus, lower KIR inhibition could allow unrestrained NK cell activation resulting in immune-mediated pathology in the brain. This would be consistent with the hypothesis that mechanisms that prevent initial *P. falciparum* infection and those that prevent severe disease are distinct and may have a balancing effect on the maintenance of different KIR and HLA ligands in malaria endemic populations.

The results of our study also underscore that case-control studies of severe vs. non-severe malaria have important limitations as a framework to identify immune susceptibility genes. Because such comparisons are restricted to individuals in whom parasitemia is already established, they cannot identify genetic factors influencing host restriction at the pre-erythrocytic stages of infection. Moreover, the development of severe malaria is likely to be influenced by genetic factors governing inflammation, as well as non-genetic factors such as socio-economic status and access to care. We believe that additional genetic determinants of malaria may be uncovered through analytic methods that incorporate rigorous longitudinal assessments of both parasite infection and symptoms, enabling evaluation of immune protection at varying stages of malaria: from initial infection and establishment of parasitemia, to development of symptomatic or even severe malaria.

While it is well established that KIRs influence the immune response to and ultimate outcome of numerous viral infections, their impact on the control of non-viral infections has been less certain. Our finding that inhibitory KIR ligands increase the risk of *P. falciparum* parasitemia indicates that the influence of KIR on host immunity extends to parasitic infections. Moreover, our findings suggest an important role for the cellular immune response in restricting *P. falciparum* infection, perhaps during the earliest stages of infection, before blood-stage parasitemia is established. These findings represent a novel genetic determinant of malaria susceptibility and further our understanding of antimalarial immunity with implications for vaccine design.

## Methods

### Study Approval

This study was granted ethical approval from the Makerere University School of Medicine Research and Ethics Committee, the London School of Hygiene and Tropical Medicine Ethics Committee, the University of California, San Francisco, Committee on Human Research and the Uganda National Council for Science and Technology. All participants and/or their parents/guardians gave written informed consent for their household upon enrollment.

### Clinical and parasitological outcomes

Data were collected from Ugandan subjects enrolled into three parallel longitudinal cohorts conducted as part of the East Africa International Center of Excellence for Malaria Research, at sites with a range of malaria transmission intensity. Nagongera, in Tororo district, is a high-transmission, rural area in southeastern Uganda. Kihihi, in Kanungu district, is a rural area in southwestern Uganda with moderate transmission. Walukuba, in Jinja district, is a relatively low-transmission, peri-urban area near Lake Victoria. Households in the three sites were enumerated and approximately 100 households per site were randomly selected. All children aged 6 months to 10 years and one adult caregiver per household who met eligibility criteria (usually a female) were invited to participate between August and October 2011. Children aged out of the cohort when they reached 11 years of age. Participants were followed through June 2016; visits after December 31, 2014 from Tororo were excluded from this analysis because transmission changed dramatically at this site after intensive indoor residual spraying campaigns were started. Analytic follow-up time was a median of 55 months in Jinja, 57 months in Kanungu, and 39 months in Tororo. Further detail on the cohort is available from Kamya et al. (Kamya et al., 2015).

Clinical data were obtained via both active and passive follow-up. All participants visited the clinic quarterly to obtain thick blood smears. In addition, caregivers were asked to bring their children to study clinics to receive care for any illness free of charge. Thick blood smears were performed on all children who reported fever in the previous 24 hours or who were febrile (>38.0°C) at the sick visit. Parasitemia was determined based on microscopy and parasite density was calculated as previously described (Rodriguez-Barraquer et al., 2018). A more sensitive measure of parasitemia (loop-mediated isothermal amplification—LAMP) was performed on samples negative by microscopy at routine visits throughout follow-up for children. However, LAMP was not conducted consistently on samples from adults at all sites throughout the study duration and thus was not included in primary analyses. Those with symptomatic malaria were treated with the first line treatment in Uganda, artemether-lumefantrine. Those with complicated or recurrent malaria (occurring within 14 days of last therapy) were treated with quinine (Rodriguez-Barraquer et al., 2018).

Entomologic data were collected monthly at all study households. Mosquitoes were collected using miniature CDC light traps and tested for sporozoites using an ELISA technique, as previously described (Kilama et al., 2014). For each household an annual entomological inoculation rate (EIR) was calculated as the product of the of the yearly household human biting rate (geometric mean of female Anopheles mosquitoes caught in a household per day) and the site sporozoite rate (average proportion of mosquitos that tested positive for *P. falciparum* in each site), as previously reported (Rodriguez-Barraquer et al., 2018).

### HLA and KIR Genotyping

HLA genotyping was performed using a targeted next generation sequencing (NGS) method. Briefly, locus-specific primers were used to amplify a total of 23 polymorphic exons of HLA-A, B, C (exons 1 to 4), DPA1 (exon 2), DPB1 (exons 2, 3), DQA1 (exon 2), DQB1 (exons 2, 3), DRB1 (exons 2, 3), and DRB3, 4, 5 (exon 2) genes with Fluidigm Access Array (Fluidigm Singapore PTE Ltd, Singapore). The 23 Fluidigm PCR amplicons were pooled and subjected to sequencing on an illumina MiSeq sequencer (Illumina, San Diego, CA). HLA alleles and genotypes were called using the Omixon HLA Explore (beta version) software (Omixon, Budapest, Hungary).

KIR genotyping for the presence or absence of each KIR gene was conducted by PCR with sequence-specific priming (PCR-SSP) as described previously (Martin and Carrington, 2008), with some modifications. Each PCR was conducted in a volume of 5 ul using 5 ng genomic DNA and SYBR Green PCR Master Mix with Platinum Taq (Invitrogen). Presence and absence of specific PCR products was detected by melting curve analysis on the 7900 Real-Time PCR System (Applied Biosystems).

### HLA-C Expression Levels

HLA-C expression levels were imputed using estimates based on previously published quantitative measurements of HLA-C allotypes (Apps et al., 2013).

### Statistics

We analyzed the association of KIR and their HLA ligands with five different outcomes related to malaria immunity:

1. Parasite prevalence: Prevalence of at least one symptomatic or asymptomatic parasite positive visit per quarter (by microscopy).
2. Annual malaria incidence rate: Number of symptomatic malaria episodes per person-year. Symptomatic malaria was defined as children with a tympanic temperature >38°C at time of presentation to clinic plus parasitemia or those with a self-reported history of fever in past 24 hours plus parasitemia.
3. Parasite density: Log parasite density measured during visit (in analyses, stratified by two visit types: routine quarterly parasite-positive visits and symptomatic malaria visits).
4. Probability of symptoms if infected: Whether a participant was febrile if they were parasitemic. (Episodes of asymptomatic parasitemia where fever developed within 7 days and repeat episodes of febrile parasitemia within 7 days of initial episode were excluded so that each malarial episode was only counted once.)
5. Temperature at parasitemic visits: Objective temperature at parasitemic visits, conditional on parasite density.

First, we assessed the effect of each KIR individually on these outcomes. Next, we explored KIR-HLA compound genotypes as follows. HLA ligand group definitions are in Supplemental Table 1. KIR2DL1, which binds HLA-C2, was present in nearly all subjects (98.6%). KIR2DL2 and KIR2DL3 segregate as alleles of the same locus and have the same HLA ligand, HLA-C1; virtually everyone in this sample has KIR2DL2 and/or KIR2DL3 (99.3%). We examined the effect of having at least one copy of HLA-C2 among those with KIR2DL1 and of having at least one copy of HLA-C1 among those with KIR2DL2 and/or KIR2DL3. Building on this, we investigated HLA-C1/C2 zygosity in the entire sample to determine the effect of multiple copies of each ligand. We then assessed the effect of a joint variable: no HLA-C1 ligand (i.e. HLA-C2 homozygous), HLA-C1 with KIR2DL2 only, HLA-C1 with KIR2DL2 and KIR2DL3, or HLA-C1 with KIR2DL3 only. We repeated this analysis among those with no KIR2DS2 (for the outcome of parasite prevalence only). Another inhibitory KIR, KIR3DL1, was also nearly ubiquitous in the sample (98.7%). We again considered the effect of its ligand, HLA-Bw4, among those with the receptor. Given that both HLA-C2 and Bw4 affected parasite prevalence in the same direction, we assessed the combined genotype of HLA-C2 and HLA-Bw4 (neither HLA-C2 nor HLA-Bw4, only HLA-C2, only HLA-Bw4, or both HLA-C2 and HLA-Bw4). Finally, we looked at the level of HLA-C expression (normalized as a z-score).

To estimate these effects, we used multi-level models with random effects at the individual- and household-levels. We used logistic regression for parasite prevalence and probability of symptoms if infected. We used Poisson models for malaria incidence. We used linear models for log parasite density, as well as temperature. All models were adjusted for site, household log entomological inoculation rate, and sex. Models included a continuous linear term for age, a binary indicator of age group (child vs. adult), and an interaction between the two to allow for effects to vary between age groups, because the study did not enroll participants between the ages of 11 and 17. Models for temperature also controlled for log parasite density at that visit.

In summary, the models followed this general form (an example model for parasite prevalence shown):

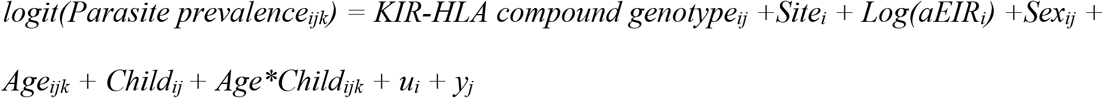

where i indicates households, j indicates individuals, and k indicates specific visits. For example, age_ijk_ denotes the age of child j from household i during visit k. As aEIR_i_ represents the average annual EIR recorded for household i (time-invariant), we assume relatively stable transmission intensity over the course of study follow-up.

We used Monte Carlo permutation tests to calculate exact p-values. We permuted exposure variables 10,000 times and calculated the two-sided p-value as the number of permutations that yielded coefficients greater than or equal to the absolute value of the observed coefficient divided by the number of models that converged (>99%) using the Stata ritest package version 1.1.4 (Heß, 2017). For each outcome, we controlled for multiple testing separately using the false discovery rate (FDR) approach with the Stata qqvalue package (Newson, 2010). All analysis was done in Stata 15.1 (StataCorp, College Station, TX).

Finally, we performed the following sensitivity analyses for all models with the outcome of parasite prevalence: stratified by site, among children only, and among sickle cell wild type. We also estimated models using a measure of parasitemia defined as positive if a subject were positive either by microscopy or by LAMP among children for the entire follow-up period and among children and adults for the period LAMP data were available for both. Controlling for ethnicity (Bantu/non-Bantu) based on the language of their consent form did not improve model fit as compared to controlling for site because these two variables were collinear.

## Data Availability

All of the clinical and parasitological data is available at the web address below.

https://clinepidb.org/ce/app/record/dataset/DS_0ad509829e

## Author Contributions

JCD, PCC, IRB and MEF designed experimental questions. IRB and JCD created statistical analysis plan and JCD performed all statistical analyses. PCC, MEF and JCD prepared original manuscript. GD, MK, EA and JR generated the clinical data set. MM and MC performed KIR and HLA genotyping. GN and MV validated statistical analyses. All authors contributed to review and editing of manuscript.

## Acknowledgments

We are grateful to all the parents and guardians for kindly giving their consent, and to the study participants for their cooperation. We thank all the members of the study team for their tireless effort and excellent work. We thank Noam Teyssier for generously sharing his computing expertise and time. Support for this work was provided by the National Institute of Allergy and Infectious Diseases (R01AI093615 and K24AI113002 to M.E.F., U19AI089674 to M.E.F., G.D. and M.K) and Henry Wheeler Center for Emerging and Neglected Diseases (to P.C.C.). In addition, this project has been funded in part with federal funds from the Frederick National Laboratory for Cancer Research, under Contract No. HHSN261200800001E. The content of this publication does not necessarily reflect the views or policies of the Department of Health and Human Services, nor does mention of trade names, commercial products, or organizations imply endorsement by the U.S. Government. This Research was supported in part by the Intramural Research Program of the NIH, Frederick National Lab, Center for Cancer Research.

## Notes

### Competing Interest Statement

The authors have declared no competing interest.

